# Towards fast, routine blood sample quality evaluation by Probe Electrospray Ionization (PESI) metabolomics

**DOI:** 10.1101/2021.04.18.21254782

**Authors:** Natalie Bordag, Elmar Zügner, Pablo López-García, Selina Kofler, Martina Tomberger, Abdullah Al-Baghdadi, Jessica Schweiger, Yasemin Erdem, Christoph Magnes, Saiki Hidekazu, Wolfgang Wadsak, Björn-Thoralf Erxleben, Barbara Prietl

## Abstract

PESI-MS enables with its greatly simplified handling and fast result delivery the application field for high-throughput use in routine settings. In health care and research, pre-analytical errors often remain undetected and disrupt diagnosis, treatment, clinical studies and biomarker validations incurring high costs. This proof-of-principle study investigates the suitability of PESI-MS for robust, routine sample quality evaluation.

One of the most common pre-analytical quality issues in blood sampling are prolonged transportations times from bedside to laboratory promptly changing the metabolome. Here, human blood (n=50) was processed immediately or with a time delay of 3 h. The developed sample preparation method delivers ready-to-measure extracts in <8 min. PESI-MS spectra were measured in both ionization modes in 2 min from as little as 2 µl plasma allowing 3 replicate measurements. The mass spectra contained 1200 stable features covering a broad chemical space covering major metabolic classes (e.g. fatty acids, lysolipids, lipids). The time delay of 3 h was predictable by using 18 features with AUC > 0.95 with various machine learning and was robust against loss of single features.

Our results serve as first proof of principle for the unique advantages of PESI-MS in sample quality assessments. The results pave the way towards a fully automated, cost-efficient, user-friendly, robust and fast quality assessment of human blood samples from minimal sample amounts.

**Graphical abstract:** 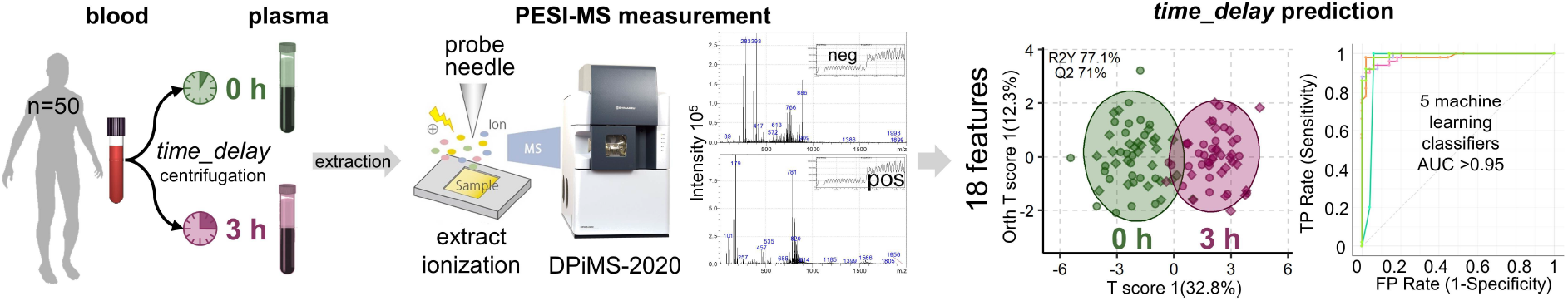

## INTRODUCTION

Mass-spectrometry (MS) is a sensitive, valuable analytical tool in different fields and substantially advanced metabolomics in the last decade. Metabolomics is defined by measuring small molecules (<2000 Da) which reflect the biological phenotype and allow mechanistic insights, identification of new therapeutic targets or development of diagnostic biomarkers. Typically, MS-based metabolomics relies on liquid or gas chromatography coupled to high resolution mass spectrometers to increase specificity, sensitivity and chemical coverage(Sands et al. 2021). Such high-quality metabolomics set-ups need extensive expert knowledge, have long measurement times, produce large amounts of data, require complicated analyses for successful research and can struggle with large sample numbers (Segers et al. 2019).

In contrast, PESI-MS omits chromatography by directly capturing tiny sample amounts with a needle tip vertically dipping into the sample. Electrospray is generated in ambient conditions by applying a high voltage to the needle (Hiraoka et al. 2007). This simplifies handling, shortens measurement times to minutes and allows previously impossible application scenarios (Hiraoka et al. 2020; Saha, Mandal, and Hiraoka 2013; Sakamoto et al. 2018), especially where robust, routine and high-throughput measurements are needed. Sample quality control is one such setting where metabolomics has shown great scientific promise but has not yet entered routine application.

High-quality biospecimens are crucial for reproducible results in *omics or biomedical research, for correct clinical routine diagnostics and for successful biobanking. Blood plasma is easily accessible and the most common biospecimen in research studies, biobanks and health care (Medina et al. 2020), reflecting the persons systemic physiological status. The processing of blood to plasma remains prone to various pre-analytical variations and errors, despite routine use and established standard operating procedures (SOP). Pre-analytical issues occur in as many as 5.5 % of in-patient routine diagnostics tests (Salvagno et al. 2008) and account for 46-68.2% of diagnostic errors (Plebani 2006). There are many different pre-analytical issues such as blood to plasma processing time delays, hemolysis, micro-clotting, under- or overfilling of sample tubes, contaminations during plasma or serum separation, and freeze-thaw cycles (Lippi et al. 2012). Time delays of blood to plasma processing occur often, e.g. when samples have to be transported from patient-site to laboratory. Such delays affect sample quality markedly and can be well detected by NMR and MS-based metabolomics (Ghini et al. 2019; Kamlage et al. 2014, 2018; Lehmann 2015; Lippi et al. 2012; Wagner-Golbs et al. 2019; Yin et al. 2013).

Despite various benefits, the use of metabolomics for quality control has not yet become an affordable, efficient and easily usable technique for routine clinical, research or biobanking. Instead, single pre-analytical issues are tested with specific methods in routine clinical diagnostic, which depend on a prior knowledge of biological processes (Hainaut et al. 2017). These single methods are still not comprehensive enough to cover the most frequent pre-analytical error sources in one swift measurement.

This complex problem could potentially be solved by exploiting the unique advantages of PESI-MS, which is able to deliver full mass spectra within minutes. In our set-up PESI was coupled to a single-quad mass spectrometer, purposely omitting more sophisticated detectors in order to (1) decrease instrument acquisition and maintenance costs, (2) increase the instruments robustness and operating usability for personnel without extensive MS expertise, and (3) decrease measurement time per sample. In this proof-of-principle study we concentrated on one of the most common pre-analytical issues, the time delay of blood to plasma processing. Our aim was to investigate if PESI-MS is able to robustly classify time delays of 3 h from immediately processed sample.

## MATERIALS & METHODS

### Study Design

The study was conducted in adherence to the Declaration of Helsinki and was reviewed by the ethical committee of the Medical University of Graz, Austria (31-116 ex 18/19, 16.01.2019). Prior to commencement of study activities, a written informed consent was obtained from all participants.

The 50 volunteers (24 female, 26 male) consisted of two subgroups, homogeneous and heterogeneous. The homogenous (n=20) was selected to reduce biological and metabolic variation. The heterogeneous group was inversely selected to increase biological and metabolic variation; an overview of inclusion criteria is reported in Table 1.

**Table 1:** Inclusion criteria homogeneous and heterogeneous group.

|  | Homogeneous (n=20) | Heterogeneous (n=30) |
| --- | --- | --- |
| **gender** | male/female | male/female/diverse |
| **age** | 20-30 | 18-90 |
| **BMI** | 18.5-25 kg/m<sup>2</sup> | ≥ 18.5 kg/m<sup>2</sup> |
| **substance abuse** | drug abuse abstinent (>1 year pre-trial)<br>nicotine ≤1 cigarette/week<br>alcohol ≤7 unit/week | at least one metabolism impacting specialty or condition<br>(e.g. special diet, ethnicity, chronic disease, substance abuse, ...) |
| Health status | healthy |  |

Exclusion criteria for all volunteers were any severe acute, or severe chronic diseases, known anemia, blood or plasma donation within the last month, pregnancy, breastfeeding, intention of becoming pregnant or not using adequate contraception, mental incapacity, unwillingness or language barriers precluding adequate understanding or co-operation as well as any other condition interfering with the volunteer’s safety.

Additional exclusion criteria for the homogeneous group were any acute or chronic disease, surgery within the last 3 months, hormonal contraception or *in vitro* fertilization (IVF) treatment in the last three months, medication with heparin, nonsteroidal or steroidal anti-inflammatory drugs in the last ten days, medication with anti-histamines or selective serotonin reuptake inhibitors in the last four weeks and any special diet form (e.g. vegan, vegetarian, gluten-free, malabsorption specific diets, raw, intermittent-fasting, ketogenic…). These exclusion criteria of the homogeneous group were inversely inclusion criteria for the heterogeneous group.

All samples were collected within 3 weeks in winter. Detailed volunteers’ characteristics can be found in Suppl. data 1, the study protocol with the informed consent is available under https://www.drks.de DRKS-ID: DRKS00024807.

### Sample collection and plasma preparation

Blood was collected in the morning between 08-10 a.m. in the homogenous group (after overnight fast) and between 08-12 a.m. (MESZ) in the heterogeneous group (without fasting). Collection was done with a 21G butterfly needle while sitting. Venipuncture was performed maximum once per arm. The tourniquet was released after 1 min and blood collection was achieved in 4 to 9 min. The first tube was discarded and each other tube was immediately inverted gently 3 times.

For the plasma preparation delay of 3 h the K_3_EDTA tubes (5 mL) were kept upright on the benchtop at room temperature (climate-controlled laboratory 20.6 to 26.2°C) until centrifugation with 2200 g for 10 min at 4°C. The plasma supernatant was aliquoted into standard protein low-bind Eppendorf tubes and aliquots were flash frozen on dry ice, transported on dry ice and stored at -80°C until measurement. Samples without *time_delay* (termed 0 h) were flash frozen within 12 to 23 min after end of phlebotomy.

### Clinical routine hematology

Tubes for clinical routine hematology were transported for 10-15 min in isolated boxes (20-25°C) within 2.5 h after phlebotomy. Hematology measurement was performed within 4 h at the Clinical Institute of Medical and Chemical Laboratory Diagnostics, Medical University of Graz (Tomin et al. 2020). Hematological measurements were carried out on Sysmex Blood counters of XN or XE series (Sysmex Co., Japan) by flow cytometry. All molecular analyses were performed with a COBAS 8000 (Roche Diagnostics, Switzerland): ions by indirect potentiometry; alanine, aspartate aminotransferase, glucose, creatinine and cholesterol by spectrophotometry; troponin T and thyroid-stimulating hormone by electro-chemiluminescence immunoassay (ECLIA) and C-reactive protein (CRP) by immunological turbidity.

### PESI-MS Measurement

Plasma aliquots were thawed in a water ice bath for 15 min. 10 µl of each plasma sample aliquot was pooled for quality control (QC) and split into 20 µl aliquots. From each sample, QC aliquot and blank a volume of 20 µl was precipitated with 380 µL extraction solution to a final concentration of 10 mM NH_4_Ac, 70% MeOH and 5% DMSO. Precipitates were frozen at -80°C until measurement within 3 h after thawing of samples. The measurement was split into 3 batches. Blanks and QC were measured repeatedly throughout the sequence (see Suppl Data 1 for measurement sequence). Precipitated extracts were thawed in 1 - 2 h (measurement time) sub-batches in water ice baths and precipitates were removed by centrifugation for 5 min at 4°C with 12.000 rpm.

Supernatants were kept in a water ice bath until measured with a PESI-MS device with LabSolutions 5.93 (DPiMS-2020, Shimadzu Corporation, Kyoto, Japan). Per replicate measurement 10 µl were deposited on a sample plate. All measurements were replicated until at least two valid TIC patterns for each ionization mode were recorded. TIC patterns were defined as invalid if there was no clear TIC spiking after 30 s of the mode (refer to Fig. 1 for invalid TIC pattern example). For instrument settings refer to Table 2. For each replicate a fresh needle (silicon coated, 18529A1, Shimadzu Corporation, Kyoto, Japan) and sample plate (11A9722418115OMS, Shimadzu Corporation, Kyoto, Japan) was used.

**Table 2:** Instrument settings for the PESI measurement.

| IONIZATION |  |  |
| --- | --- | --- |
| Needle position | -37 mm |  |
| Outage time | 200 ms |  |
| SAMPLE TAKE |  |  |
| Needle position | -46 mm |  |
| Outage time | 50 ms |  |
| Speed | 250 mm/s |  |
| MEASUREMENTS |  |  |
| voltage segment | Electric potential | Time |
| 1 - Corona | +4.00 kV | 3.6 s |
| 2 – neg mode | -4.25 kV | 30.0 s |
| 3 – Corona | -4.50 kV | 3.6 s |
| 4 – pos mode | +2.75 kV | 30.0 s |
| m/z range | 10-2000 m/z |  |
| Scan speed | 5000 u/s |  |

**Fig. 1:**
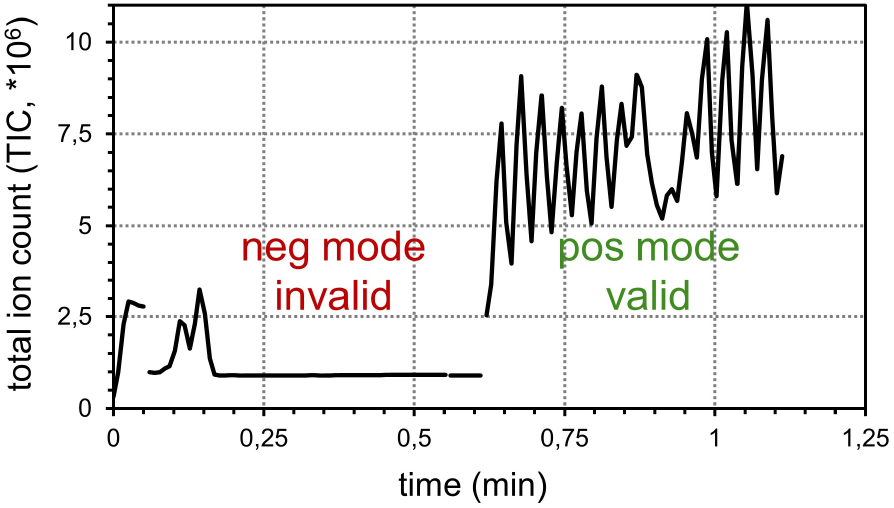
Example graph of a PESI measurement diagram. The negative mode (neg mode) TIC pattern is invalid, the positive mode (pos mode) is valid.

## Data extraction and pre-processing

Mass spectra of all measurements were exported in JCAMP-DX format (Lampen et al. 1994) for each voltage segment. Features were extracted with eMSTAT 1.0 (Shimadzu Corporation, Kyoto, Japan) separate per ionization mode but for all measurements at once by binning with a m/z tolerance of 0.75Da. The intensity threshold was 0.1% for neg mode (voltage segment 2) and 0.01% for pos mode (voltage segment 4). Resulting intensities for each sample (in rows) and each m/z binned feature (in columns) were copied to Microsoft Excel (2013), combining both ionization modes for each measurement resulting in 4702 features (refer to Table 3 for example of data structure).

**Table 3:**
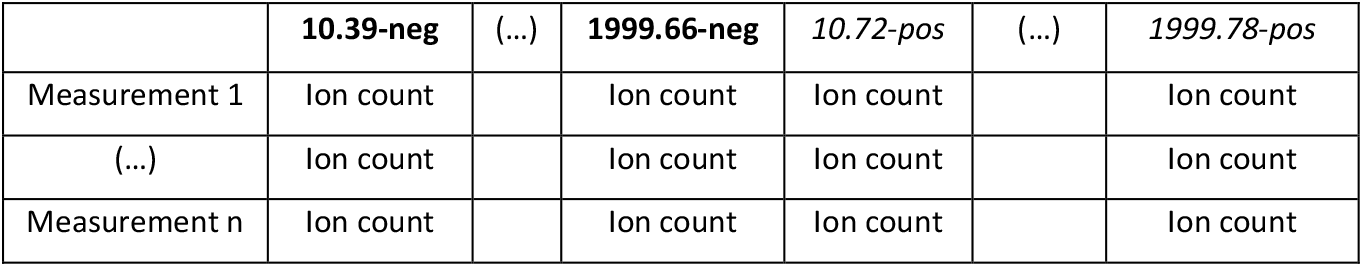
Shematic of eMSTAT intensity export structure, with samples in rows and features (name designation by m/z and ion mode).

Further data extraction and pre-processing was performed with Tibco Spotfire (11.1.0). All invalid single modes were excluded from any further calculations. LOG data was created by averaging all valid replicate measurements of a sample and log_10_-transformation. LOG_QCBC data was created by applying a metabolomics workflow including trimming, batch correction and filtering of low-quality features (refer to Table 4 for criteria and Supplementary Figure 3 for example schematics). Corrected and filtered data was averaged for valid replicate measurements, QC and blanks were excluded and all values were log_10_-transformed. After filtering 1200 features were used for further statistical analysis (from the initially exported 4702 features) as either LOG or LOG_QCBC data.

**Table 4:** Overview of applied data filtration and batch correction steps.

| Step | based on | orientation or cut-off | Effect |
| --- | --- | --- | --- |
| **1. TIC filter** | TIC pattern | per ionization mode | excludes unstable modes of replicate measurements |
| **2. Trim** | MAD Score(Leys et al. 2013) on $\log_{10}$ | per feature & samples type (QC, sample, blank) & batch | excludes single, strong outliers setting them to NULL (= missing value) |
|  | percentage of data trimmed | <12.5% per feature | removes noisy and unstable features |
| **3. Batch correction** | median of QC in batch | per feature & batch | reduces absolute intensity differences between batches |
| **4. Filter low quality features** | average intensity in QC | per feature and mode<br>neg $>1 \cdot 10^3$, pos $>9 \cdot 10^3$ | removes background noise features with very low intensity |
|  | RSD in QC | per feature, <50% | removes noisy features with low reproducibility |
|  | missing data | per feature<br><30% in QC, <30% in samples | removes features with unstable signal to allow later use of imputation for MVA |
|  | blank load | per feature, <50% | removes background features |
| **5. Average, $\log_{10}$** | average replicates, $\log_{10}$ | per sample | combined replicates into one sample measurement, remove technical measurements, improve data distribution |

### Statistical analysis

Data visualization and statistical analysis was performed with R(R Core Team 2020) (v3.5.3, packages *stringr, dplyr, readxl, openxlsx, nlme, emmeans, ggplot2, ggpmisc, pheatmap, RColorBrewer, colorspace, dendsort, missMDA, mixOmics, MetaboAnalystR*)(Chong and Xia 2018; Pinheiro et al. 2015; Rohart et al. 2017; Wickham 2011), Tibco Spotfire (v11.1.0) and the Orange data mining toolbox (Demšar et al. 2013).

Normality and scedasticity was tested by Kolmogorov-Smirnov test with *stats::ks*.*test()* and by Bartlett test with *stats::bartlett*.*test(*), applying Benjamini-Hochberg (BH) multiple test adjustment with *stats*::*p*.*adjust(*). Hierarchical clustering analysis was performed centred and scaled to unit variance *base::scale()* over parameters and over samples. Dendrograms were calculated with standard clustering *stats::hclust(dist())* (Lance-Williams dissimilarity update with complete linkage) and sorted *dendsort::dendsort()* according to the average distance of subtrees at every merging point. Heatmaps were plotted using *pheatmap::pheatmap()*.

Principal component analysis (PCA) was performed centered and scaled with *mixOmics::pca()*. For sparse and independent PCA *mixOmics::spca()* and *::ipca(*) missing values were beforehand imputed *missMDA::imputePCA* and *missMDA::estim_ncpPCA*. Sparse projections to latent structures discriminant analysis (PLS-DA) was performed centered and scaled with *mixOmics:: plsda ()* and missing values were imputed beforehand analog as for sparse PCA.

Orthogonal PLS-DA (OPLS-DA) was performed centered and scaled to unit variance with *MetaboAnalystR::OPLSR*.*Anal* (option *scaleNorm=“AutoNorm”*) with a standard 7-fold cross validation for the classification factor *time_delay*. Model stability was additionally verified with 1000 random label permutations and models with Q^2^>50% were considered significant.

### Predictive models

To determine the suitability of PESI for discriminating samples, five common machine learning classifiers were used with standard configuration and 10-fold-cross validation (90% training, 10% test) with the Orange data mining toolbox (refer to Table 5).

**Table 5:** Evaluated classifiers and their parameters.

| Classifier | Configuration |
| --- | --- |
| Logistic Regression | Ridge (L2) regularization, C=1 |
| Tree | Binary tree, minimum 2 instances per leave, do not split subsets smaller than 5, maximum tree depth: 100, stop when majority reaches 95% |
| Random Forest | 10 trees, do not split subsets smaller than 5 |
| Support Vector Machine (SVM) | Type: SVM, C=1, epsilon=0.1, RBF Kernel (g auto), Iteration limit 100, numerical tolerance 0.001 |
| Neural Network | 100 neurons in hidden layers, ReLu activation, Adam solver, max 200 iterations, regularization alpha=0.0001 |

The influence of feature selection was investigated, since more features (1200) than samples (100) were analyzed. Feature importance was calculated for all 1200 features on both LOG and LOG_QCBC data with Orange based on five criteria (ANOVA, Gain ratio, Gini, Info.gain, *χ*^2^) for the given classes of *time_delay* {0 h, 3 h} and exported as .csv for further analysis with Tibco Spotfire. For each criterion and data type (LOG or LOG_QCBC) feature importance was ranked and plotted in descending order. Cut-offs for inclusion were set directly before importance leveled, above the “elbow” of the scree-like curve: Info.gain ≥ 0.1, Gain ratio/Gini ≥ 0.06, ANOVA ≥ 10, *χ*^2^ ≥ 6. Features were included when importance was above the cut-offs in at least one of the criteria (29 in LOG and 27 in LOG_QBCBC). Thereof 18 features overlapped and were selected for further analysis.

## RESULTS

### One-step precipitation delivers both ionization modes in one run

One of the most common pre-analytical issues is the time delay of blood to plasma processing. Blood samples were obtained from 50 volunteers, subdivided into a biologically homogeneous and heterogeneous group (refer to Fig. 2A). Routine hematological and clinical parameters showed no biological outliers when checked with unsupervised PCA and hierarchical clustering (Supplementary Figure 2), verifying suitability and inclusion of all 50 volunteers.

**Fig. 2:**
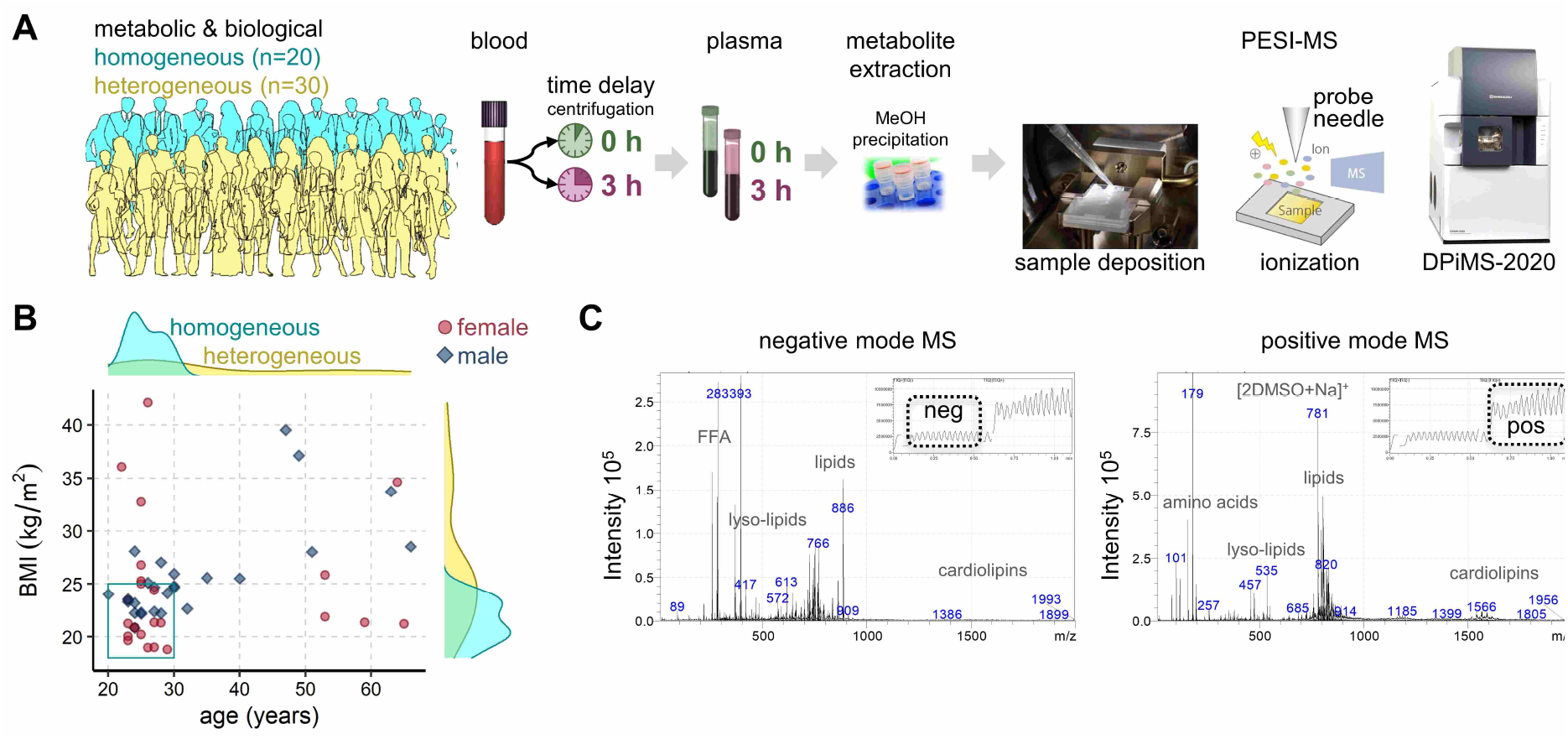
Overview of cohort and measurement workflow. **A:** In total 50 volunteers donated blood, with one sample being processed to plasma immediately *time_delay* = 0 h and another one with *time_delay* = 3 h, simulating a typical transportation time from bed-side to laboratory. Metabolites were extracted in a one-step 70% MeOH precipitation (10 mM NH_4_Ac, 5% DMSO) and 10 µl supernatant were measured with a DPiMS-2020 PESI-MS. Figures of the sample chamber, probe electrospray ionization and DPiMS-2020 table-top instrument are a courtesy of Shimadzu Corporation. **B:** Age and BMI distribution of the subgroups. The cyan box marks the age and BMI inclusion cut-offs for the homogenous group. **C:** Example full mass spectra for each ionization mode with the TIC pattern provided as inset. Possible metabolite classes based on mass ranges were added for a tentative orientation.

From each volunteer one blood sample was processed into plasma immediately (*time_delay = 0 h*), while a second sample was delayed for 3 h (*time_delay = 3 h*). Metabolites were then extracted by a simple one-step 70% MeOH precipitation with 10 mM NH_4_Ac and 5% DMSO and the diluted supernatants were measured with the PESI-MS (see Fig. 2A). Instrument settings were optimized to deliver stable results from both ionization in one run covering each the mass range from 50-2000 Da (refer to Fig. 2C). Method development details are described in the supplementary information. For easier orientation, mass ranges were tentatively marked with one possible metabolite class, which would be expected to exist in sufficient concentration in plasma extracts.

A total of 4702 features were available after combination of both modes. Low quality signals were filtered and excluded (refer to Supplementary Figure 3), retaining 1200 features for further analysis. Both modes contributed roughly half of all features (refer to Supplementary Figure 3, step 4). Feature mass ranges partially overlapped between both modes (refer to Fig. 2C). Additionally, in each mode features were found in mass ranges not detected in the other mode, e.g. amino acid masses in pos mode, FFA masses in neg mode.

Two types of data processing were used, the simple LOG and the metabolomics typical LOG_QCBC (QCBC = quality control batch corrected). For LOG data all valid replicates were averaged and data was log_10_-transformed. For LOG_QCBC data was trimmed by MAD score and batch effects were corrected by median QC (refer to Supplementary Figure 3). The technical variability ranged around 30% median RSD in QC and was lower in LOG_QCBC (26.7%) than in LOG (34.6%) data. All 1200 features were found to be sufficiently normally distributed according to Kolmogorov-Smirnov test and sufficiently homoscedastic according to the Bartlett test (Benjamini–Hochberg adjusted p-value>0.05, see Supplementary Data 1) in both data types (LOG, LOG_QCBC).

### Time delay in plasma preparation is predictable with selected features

An unsupervised PCA showed that the similarity of samples is dominated by inter-personal variability (refer to Fig. 3A). No other trends were detected within the first six components according to other biological or experimental factors (age, gender, BMI, WHR, subgroup). The batch had no detectable impact on LOG_QCBC data while in LOG data single features were impacted (refer to Supplementary Figure 3). The supervised machine learning approach OPLS-DA found no significant difference for *time_delay* 0 h vs. 3 h. Five other common machine learning approaches were used to classify *time_delay* of plasma preparation (see Fig. 3B). Performance differed notably, with three performing well with an AUC>0.8 (logistic regression, decision tree, random forest) while the two others (SVM, neural network) failed to predict *time_delay*.

**Fig. 3:**
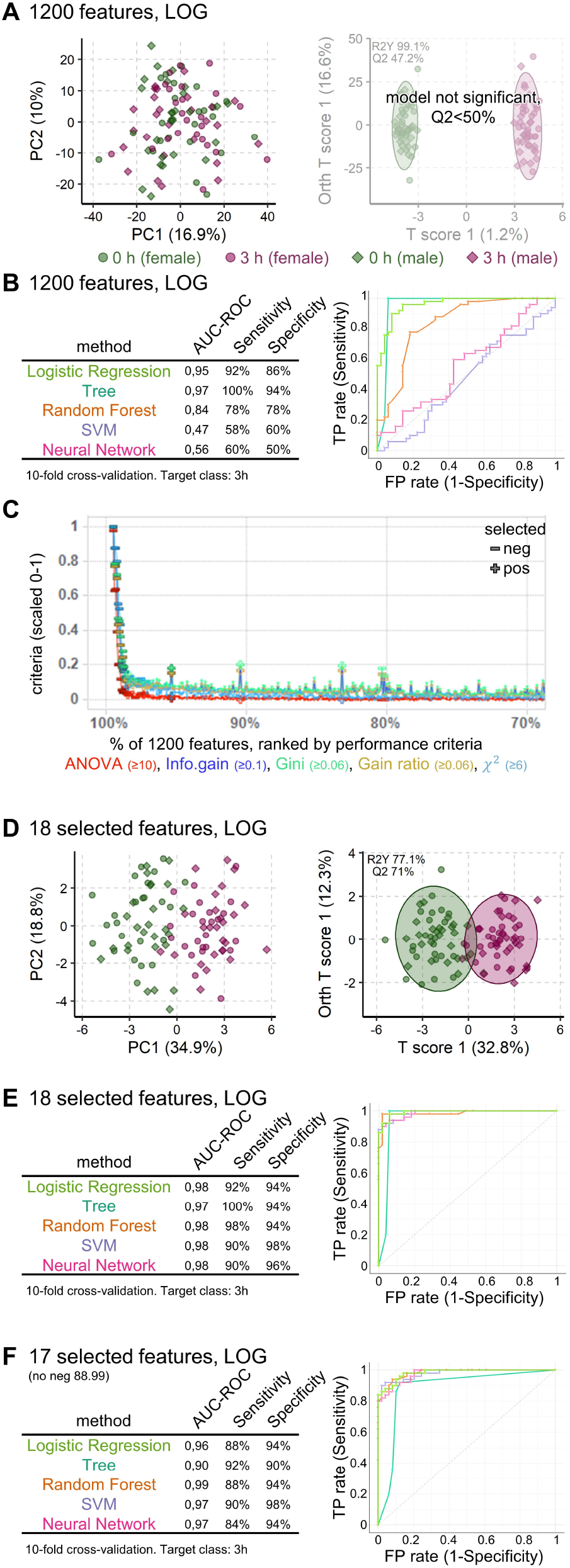
Plasma preparation delay is detectable from selected PESI features with high specificity. **A:** The unsupervised PCA (left) and supervised OPLS-DA (right) scores plot show no significant separation of the whole metabolome (1200 features, LOG data from 50 volunteers) based on *time_delay* (0 h, 3 h). **B:** From five applied machine learning approaches only three were able to predict *time_delay* (AUC>0.8) and ROC curves differed notably between the different approaches. **C:** Feature importance in each of the five test criteria are shown ranked by highest importance first. The importance in each criterion was scaled 0-1 to allow direct comparison. For clarity only top 30% of data (360 features) are shown. The importance of the other feature were all below the inclusion threshold. **D:** Both PCA and OPLS-DA scores plot show a clear and highly significant difference for *time_delay* 0 h vs. 3 h, when based on the selected 18 features (LOG). **E:** All five applied machine learning algorithms delivered predictions of *time_delay* (AUV>0.95) with no false negatives and very similar ROC curves.

The homogeneous group was selected to reduce biological variability to possibly improve detection of sample quality biomarkers. However, PCA and OPLS-DA with the homogeneous group alone with all 1200 features resulted in similar unspecific findings (refer to Supplementary Figure 5).

As a next step, sparse and independent multivariate methods were tested. Unsupervised sparse and independent PCA both failed to improve group separation (refer to Supplementary Figure 6A, B). Supervised sparse PSL-DA was able to detect a significant group separation in the first component, when tuned to retain ten features. Prediction performance for supervised sparse PSL-DA was excellent with an AUC>0.95 (refer to Supplementary Figure 6C, D, E).

Consequently, a more extended feature selection was used to exclude the bulk of noisy or unspecific features to further reduce noise. The selection was based on five diverse criteria (ANOVA, Gain ratio, Gini, Info.gain, *χ*^2^) to avoid overlooking possibly well performing features when concentrating on one selection criteria. The criteria were applied in an univariate manner, calculated for each feature independently. Features were selected if any of the five criteria showed sufficient importance in LOG and LOG_QCBC data via the aforementioned criteria,resulting in 18 selected features (see Fig. 3C). These 18 selected features encompassed all ten features from the first component of the sparse PLS-DA.

The 18 selected features were used in the same unsupervised and supervised analyses (refer to Fig. 3D, E). The PCA exhibits a clear group separation according to *time_delay*, which is found to be highly significant with OPLS-DA (Q^2^>70%, refer to Fig. 3D). All five machine learning approaches performed consistently excellent with an AUC>0.95 (refer to Fig. 3E).

The same analyses were repeated with LOG_QCBC data to assess how improved data quality with reduced technical variability impacts predictive performance. Overall performance was similar or better, for example OPLS-DA became significant even with all 1200 features (refer to Supplementary Figure 4).

### Selected features form a robust pattern for prediction of time delay in plasma processing

The selected 18 features were investigated in more detail using a heatmap and single scatter plots (refer to Fig. 4 LOG and Supplementary Figure 7 LOG_QCBC). Most features (16) increased with *time_delay* and only two features showed a decrease (neg 812.1, neg 787.84). The best performing feature neg 88.99 stems with high probability from lactate. This feature alone would be able to predict *time_delay* of 3 h. Predictions with the remaining 17 features without the top driver neg 88.99 also performed excellently (see Fig. 3F). Only one feature stemmed from the pos mode (pos 974.8), all other features were negative ions. The neg features consistently showed larger differences with *time_delay*.

**Fig. 4:**
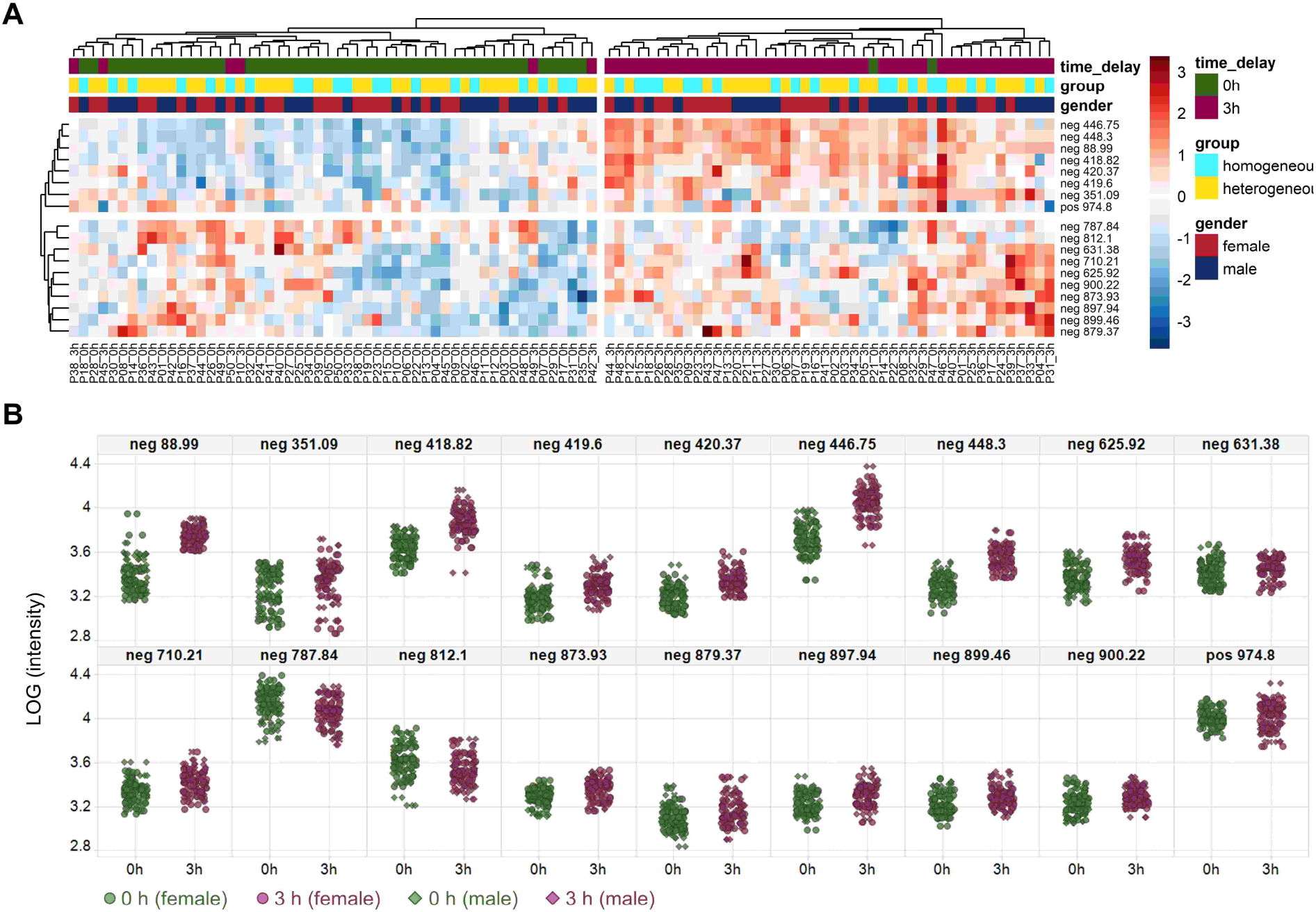
Heatmaps and plots of the 18 selected features in LOG data. **A:** Heatmaps with hierarchical clustering show the difference induced by *time_delay* in the 18 most important features. There are no systematic differences between study subgroups or genders. **B:** Scatter plots showing that from the 18 features, most (16) increased after the 3 h *time_delay* while only two feature levels decreased.

## DISCUSSION

In this study we obtained blood plasma samples from 50 volunteers, developed a measurement method covering both ionization modes and used advanced machine learning analysis to conclusively show the potential of PESI-MS for routine sample-quality determination.

From each volunteer a blood sample was processed into plasma immediately (*time_delay = 0 h*), while processing of the second sample was delayed for 3 h (*time_delay = 3 h*) to simulate a typical transportation time from bedside to clinical laboratory. The 3 h time delay was selected as show case because most routine clinical read-outs remain valid while sensitive analytes already show considerably degradation, e.g. in metabolomics (Kamlage et al. 2014) or proteomics (Rai and Vitzthum 2006).

The preparation of plasma samples for measurements should be fast, simple, cost-efficient, use minimal sample amounts and be compatible with full automation for high-throughput. A more detailed description of the method development can be found in the supplementary information. In short, measurements directly from plasma or simple dilutions gave insufficient signal quality (signal density, reproducibility, ionization mode coverage). However, a simple one-step 70% MeOH precipitation with 10 mM NH_4_Ac and 5% DMSO was found to deliver reproducible results in both ionization modes (neg and pos). The whole sample preparation can be performed manually with standard laboratory equipment in less than 8 min total time, including the 5 min centrifugation step. This time can be reduced to <1 min by switching to filtration. For the 2 min PESI-MS measurement 10 µl extract sufficed, so that 2 µl plasma enabled three replicates with the applied 1:20 dilution during precipitation. In this study 20 µl plasma were standardly used because enough sample material was available and the higher volumes were more convenient for manual handling.

Data analysis was primarily focused on LOG data because the aim is a future routine application, which would strongly profit from not relaying on QCs, blanks and batch effect corrections. However, initial search for sample quality biomarkers with all 1200 features failed with unsupervised PCA, supervised OPLS-DA and five common machine learning approaches (refer to Fig. 3A, B). The unsupervised PCA scores plot was dominated by inter-individual variability. The performance discrepancies between the five machine learning approaches also indicate that data variability was too high.

The selected simple and cost-efficient instrument has many advantages as listed in the introduction, though the unavoidable trade-offs are notably noisier signals with extensively mixed isobaric overlaps. Noise and overlapping signals decrease statistical power and thus chances to detect sample quality specific biomarkers. Therefore, the cohort was subdivided into a homogenous subgroup (n=20, refer to Fig. 2A) with reduced biological and metabolic variability by including specifically healthy, young, non-obese volunteers (see Fig. 2B). The homogeneous group was also sampled in a short 2 h morning time window after an overnight fast (12 h) within 3 weeks to exclude seasonal influences, according to typical recommendations of metabolomics or proteomics studies (Kamlage et al. 2014). In contrast, the heterogeneous subgroup (n=30) aimed for high variability by diversifying the volunteers age, weight, life-style and health-state. The heterogeneous group was designed to assess the robustness of an identified biomarker against real-life human variability.

However, PCA and OPLS-DA of the homogeneous group alone failed with all 1200 features and resulted in similar unspecific predictions (refer to Supplementary Figure 5), which demonstrates that biological variability is not the decisive factor in this setting. Consequently, future studies could omit the cumbersome step of subdividing cohorts and reducing biological variability when searching for plasma quality biomarkers.

We further investigated the aspect that multivariate methods can be susceptible to too many noisy or unspecific features, drowning out signals of a few highly significant features. Here the term noisy refers to high technical variability and the term unspecific refers to not reflecting changes for the factor of interest (*time_delay*). Therefore, sparse and independent methods provided in the mixOmics package(Rohart et al. 2017) were tested. Sparse methods include a feature selection by ℓ1 regularization directly implemented into the optimization criterion. The independent methods use an independent component analysis for de-noising loadings vectors to obtain independent PCA components. Unsupervised sparse and independent PCA both failed to provide additional insights (refer to Supplementary Figure 6A, B). In contrast, the supervised sparse PLS-DA was able to detect significant differences for 3 h *time_delay*, when set to retain ten features in the first component.

This indicates that the high feature numbers rather than biological variability hinder detection of sample quality specific biomarkers. Although the sparse PLS-DA detected 10 highly predictive features (AUC>0.95), the selection to retain 10 features was arbitrary and could miss further predictive features. Accordingly, features were filtered to signals of interest using five diverse, univariate criteria arriving at 18 features of interest (see Fig. 3C). These 18 features encompassed all ten features identified by sparse PLS-DA.

Analogue analyses were repeated with unsupervised and supervised PCA, OPLS-DA and five machine learning algorithms. All methods clearly demonstrated impact of *time_delay* when concentrated on the 18 features of interest, instead of using all available 1200 features (see Fig. 3D,E). All machine learning approaches were able to predict *time_delay* with an as excellent AUC>0.95 as sparse PLS-DA.

In general, others reported a considerably larger percentage of impacted metabolites by time delays in plasma processing (Ghini et al. 2019; Kamlage et al. 2014, 2018). For example, Kamlage et al. (Kamlage et al. 2014) reported that 10% of all metabolites increased and 12% decreased after a time delay of 2 h at room temperature. Here, only 18 of 1200 features were changed, corresponding to 1.5%. One likely reason is the higher technical variability with a median RSD in QC of ∼30% when compared to 5-15% in more sophisticated metabolomics methods.

Hence, we compared if lower technical variability could improve detection of predictive biomarkers by also analyzing batch corrected, trimmed LOG_QCBC data. LOG_QCBC data reduced technical variability by 7.9 percentage points (median RSD) when compared to LOG data. Results with LOG_QCBC data were similar to LOG results with slightly improved performance, for example in OPSL-DA (all and selected features). A reduction of technical variability will be beneficial for future robust high-throughput application, but the used batch correction and trimming is not compatible with routine and stand-alone measurements. Accordingly the use of internal standards should be investigated, where RSDs below 20% were reported with PESI-MS (Saha, Mandal, and Hiraoka 2013). Additionally, automatisation and optimization of sample preparation will further decrease technical variability.

From the selected 18 features 16 decreased, while only two increased; 17 were from neg mode, while only one was from pos mode (refer to Fig. 4 LOG and Supplementary Figure 7 LOG_QCBC). The use of one ionization mode would halve measurement time, doubling throughput and will thus be very beneficial for future routine applications. Nevertheless, we would not recommend abandoning the pos mode at this stage of development. The combination of both modes notably expands the covered chemical space. This could prove crucial when expanding search of sample quality biomarkers into other pre-analytical factors such as freeze thaw cycles, hemolysis, micro-clotting, long-term storage or into other sample types such as serum or urine.

The feature masses were manually checked in the Human Metabolome Database (HMDB) (Wishart et al. 2018) assuming a single charged adduct to investigate from which known blood metabolite the signal could stem. All metabolite attributions are clearly speculative because a classical annotation is impossible with single quad spectra from direct ionization. All PESI-MS features will be reflecting a mixture of multiple isobaric metabolites due to the mass exactness of ±0.2 Da and the lack of chromatographic separation. Nevertheless, the tentative assignments allow some comparison to results from other studies.

Neg 418.82 could correspond to inositol 1,4,5-trisphosphate (IP3, HMDB0001498). IP3 is known to be released by erythrocytes and would be a plausible explanation for the observed increase after the 3 h time delay (Downes and Michell 1981). Several features could correspond to complex lipids: neg 631.38 to DG(36:3), neg 710.21 to PE(34:4), neg 873.93 to TG(54:8), neg 879.37 to PGP(38:3), neg 897.94 to TG(56:3) and neg 899.46 to PGP(40:7) with the abbreviation denoting the lipid type (DG – diacylglycerols, PE – phosphatidylethanolamine, TG – triacylglycerides, PGP – phosphatidylglycerolphosphate) and the numbers denoting the total chain length and number of unsaturated bonds. Increases of triacylglycerols have been reported and assumed to result from glycerophospholipid cleavage (Heins, Heil, and Withold 1995). These can be further degraded to diacylglcyerols (Hess 2014) and would plausibly explain the observed increases.

Notably, the masses of neg 418.82 and neg 419.6 are spaced by roughly 1 Da and the signal intensity of the heavier feature was lower than of the lighter feature. Possibly both features stem from the same metabolite with the lighter feature reflecting the monoisotopic peak. In such a case the patterns could be further simplified without loss of information content by excluding the isotope peak or summarizing the features.

The best performing feature neg 88.99 stems probably from lactate. Lactate is also well known to increase with time as an end product of erythrocyte driven glycolysis (Kamlage et al. 2014, 2018). The neg 88.99 performance alone would suffice for very good prediction of a 3 h time delay. Prediction was as good without the neg 88.99, showing that the suggested features form a robust pattern against single feature failures. Robustness against single feature failures is important for routine high-throughput applications reducing the need for repeated measurements. Additionally, medical conditions possibly invalidating single features would not impede a sample quality determination based on a multi-feature read-out.

One could argue that a focused single-feature method, e.g. based on lactate alone would suffice to determine a time delay before plasma processing. Many such highly sensitive methods exist or are under development, for example also from our group based on glutathione (Tomin et al. 2020). Such focused methods offer considerable advantages, ranging from easier automatization, lower cost, lower complexity, simpler transferability to other measurement platforms up to faster regulatory in vitro diagnostic (IVD) approval. However, they have limitations as they cannot cover multiple pre-analytical issues in one measurement, have to be repeated when single feature signals technically fail and can become invalid for certain medical conditions.

Our aim was to determine whether PESI-MS has the potential to determine sample quality, for which one pre-analytical issue was used as proof-of-principle. Our results demonstrate that PESI-MS spectra contain multiple robust biomarkers. Additionally, many other stable features were detected which renders detection of robust biomarkers for other pre-analytical highly likely.

### Limitations of the study

Despite PESI-MS being suitable to detect a plasma processing time delay of 3 h, only one time point was investigated allowing only a binary read-out <3h or >3h. Time series and expanded total duration would allow to determine the actually elapsed time, which is important to provide a more fine-tuned insight into data quality. Degradation process starts in blood directly after phlebotomy and while one biological read-out might become invalid after a few minutes (Tomin, Schittmayer, and Birner-Gruenberger 2020) other can remain stable over many hours.

The observed technical variability was comparatively high which decreases statistical power. Although still highly predictive features were found, a reduction of technical variability by internal standard or further sample preparation optimization will be beneficial. Additionally, other blood sample types (e.g. heparin, citrate, serum) and sample quality issues should be investigated, such as hemolysis, micro-clotting, freeze-thaw-cycles or long-term storage. In this study 50 volunteers were included, which is a limited number to reflect clinical real-life heterogeneity. Final patterns should be based on considerably more study subjects.

The lack of automatization is another major limitation towards routine and high-throughput use and could be improved by an autosampler and filtration instead of centrifugation during precipitation. However, we consciously concentrated on a proof-of-principle study in hopes to encourage further research and technical development.

## CONCLUSIONS

Our results provide a proof-of-concept that PESI-MS is a promising technology for fast and comprehensive quality control of blood samples.

The single-step MeOH precipitation delivered ready-to-measure extracts in <8 min when performed manually and could be considerably sped-up with filtration and automatization. As little as 2 µl plasma sufficed for PESI-MS spectra in both ionization modes in 2 min with 1200 stable features covering a broad chemical space. The time delay of 3 h was predictable with five common machine learning approaches based on 18 selected features with an excellent AUC > 0.95 and was robust against failure of single features. Even though for a future high-throughput application more optimization, reduction of noise and automatization are needed, our results demonstrate the unique advantages of PESI-MS. These results pave the way towards a fully automated, cost-efficient, user-friendly, robust and fast quality assessment of human blood samples from minimal sample amounts.

## Data Availability

The authors confirm that the data supporting the findings of this study are available within the article and its supplementary materials. Additional clinical data can be obtained upon request from the corresponding author.

## Acknowledgment

We thank Eva Svehlikova, Sigrid Deller, Elisabeth Langmann, Rasa Vitonyte, Robert Stefan Lipp, Amar Alikadic, Jehona Qerimi-Hyseni, Ines Mursic, Andrea Halsegger, Michael Wolf, Martina Urschitz, Ana Semonik, Martina Brunner, Rene Peter Engel, Beatrix Stroisnik, Georg Wiesnegger, Sven Miedler, Annemarie Marold and Julia Matejka from the Clinical Reseach Center (CRC) of the Medical University Graz for the excellent implementation of the clinical study and their active, always kind support and great discussions. We thank Ruth Birner-Grünberger, Matthias Schittmayer-Schantl, Tamara Tomin, Björn Thoralf Erxleben, Stephane Moreau, Christopher Titman and Takaaki Hiraoka for their excellent support, professional work, great discussions and kindness.

The samples/data used for this project have been provided by Biobank Graz, Austria.

## Author contributions

Conceptualization and supervision, NB; Data curation, NB, EZ, PLG, SK, MT, AB, JS, YE; Formal analysis and visualization, NB, PLG; Investigation, NB, EZ, SK, MT, AB, JS, YE, SH; Methodology NB, SH, PLG, EZ; Validation and writing original draft, NB, PLG, EZ; Software, NB, PLG, SH; Resources, NB, EZ; PLG, CM, SH, BTE, WW, BP; Project administration, NB, EZ, SH, BP; Funding acquisition, NB, BTE, WW, BP; Writing—Review & Editing, all authors

## Abbreviations

ACN: acetonitrile
ANOVA: analysis of variance
AUC: area under the curve
BH: Benjamini-Hochberg
BMI: body mass index
DG: diacylglycerols
DMSO: dimethyl sulfoxide
EtOH: Ethanol
FFA: free fatty acids
FP: false positive
HMDB: Human Metabolome Database
IP3: inositol 1,4,5-trisphosphate
IVD: in vitro diagnostic
IVF: in vitro fertilization
logLik: log-likelihood
MAD: median of all absolute deviations
MeOH: methanol
MS: mass spectrometry
NH_4_Ac: ammonium acetate
PCA: principal component analysis
PE: phosphatidylethanolamine
PGP: phosphatidylglycerolphosphate
OPLS-DA: orthogonal projections to latent structures discriminant analysis
PESI: probe electrospray ionization
QC: quality control
ROC: receiver operator characteristic
RSD: relative standard deviation
SVM: support vector machine
TIC: total intensity count
TG: triacylglycerides
TP: true positive
WHR: waist to hip ratio

## SUPPLEMENTARY

### Development of plasma metabolite extraction for immediate PESI-MS

The plasma precipitation method was developed to allow high-throughput use in routine laboratories aiming at: (1) low chemicals and consumable costs (<5€/sample), (2) no highly toxic or dangerous solvents necessitating non-standard safety measures, (3) using only standard laboratory equipment (e.g. vortexer, pipets, water bath, freezers, centrifuge), (4) fast work-up with a maximum time until measurement of <30 min, (5) short hands-on time with <5 min per sample, and (6) compatible with high-throughput automatization.

For ionization and reproducibility tests, normal human pooled plasma (PP) (K2 EDTA, Innovative Research Inc., IPLA-N) was used. The suitability of a preparation method was judged by: (1) information content [i.e. number of features, total intensity count (TIC) and m/z coverage], (2) ability to detect significant differences between freshly prepared PP and PP degraded for 4 h to 24 h at room temperature, (3) reproducibility (median RSD in replicates in features), (4) blank load, and (5) stability of the extract during one measurement day (tested 15 min up to 8 h at room temperature or in a water ice batch).

Prior PESI-MS work showed that signal depends on various factors: needle (thickness, surface, surface oxidation), needle immersion depth, repetition frequency, solvent type, ion strength, and applied voltage among others (Hiraoka et al. 2007; Johno et al. 2018; Lee et al. 2009; Mandal et al. 2012, 2013; Mandal, Chen, and Hiraoka 2011; Ninomiya et al. 2018; Saha, Mandal, and Hiraoka 2013; Sakamoto et al. 2018; Yoshimura et al. 2011, 2012, 2009).

Since the fundamental ionization principles are comparable between PESI and ESI, sample preparation knowledge from ESI can be transferred to PESI sample preparation to some extent. Briefly, in most ESI applications the solvents MeOH, MeOH/ACN/acetone and ACN, outperform other solvent mixtures (Sitnikov, Monnin, and Vuckovic 2016), while for PESI use of either MeOH or EtOH is more common. In ESI the solvent type has often more impact than the solvent percentage (Sitnikov, Monnin, and Vuckovic 2016). A final water content of 30-50% with an ion strength between 1–100 mM is most often optimal for spray formation and efficient analyte ionization (Silas G. Villas-Boas, Jens Nielsen, Jorn Smedsgaard, Michael A. E. Hansen 2007). Typically, volatile acids or salts are used in low concentrations such as formic acid, acetic acid or NH_4_Ac. Especially NH_4_Ac has the advantage of inducing a neutral pH allowing for positive and negative mode ionization and being soluble in up to 90% ACN(Dolan n.d.; Silas G. Villas-Boas, Jens Nielsen, Jorn Smedsgaard, Michael A. E. Hansen 2007). Others suggestion include the addition of “super-chargers” such as DMSO to improve S/N ratio(Liu et al. 2012; Valeja et al. 2010). DMSO is a very good solvent and likely contributes to signal enhancement by increased solubility. In both modes several features corresponding to possible DMSO fragments or adducts were found, with accordingly very high signals in the blanks. In each measurement the strongest signal was pos 179.1 Da which is most likely [2DMSO+Na]^+^. These strong DMSO signals could potentially serve as internal standards for normalization.

For PESI, analyte concentrations as low as 10–50 nM were found by others to yield sufficient signal(Ninomiya et al. 2018). In literature, several steps had minimal or non-beneficial effects on performance and were not tested here: solid phase extractions (Sitnikov, Monnin, and Vuckovic 2016), ultra-filtration (Nagana Gowda and Raftery 2014), acid precipitations (Bruce et al. 2009; Zhao and Xu 2010), multiple extractions, prolonged vortexing or bead-milling of plasma samples (different for tissue/cells).

According to our aims only simple preparation methods were tested, purposely omitting steps with prolonged incubations or evaporations to dryness. The factor dilution was tested from undiluted to 1:100, with dilutions around 1:20 being a good trade-off between reduction of matrix suppression and signal intensity. From the tested solvents (EtOH, MeOH, ACN, MeOH/ACN/acetone) overall TIC patterns were clearest from 70% MeOH, followed by dilutions in 50% EtOH. A strong, two-step precipitation (i.e. first adding pure solvent, than adding water) showed no additional benefit over a one-step, direct precipitation with the solvent/water mixture and was therefore not pursued further. For both MeOH and EtOH, the addition of 10 mM NH_4_Ac and 5% DMSO further stabilized and increased the TIC. Applied voltage and needle immersion depth had to be optimized for each solvent type, with e.g. an immersion depth of 46 mm for MeOH but 45 mm for EtOH. The presence of 5% DMSO allowed for 0.25 kV lower voltage, while MeOH needed 0.25 kV more in both ionizations modes compared to EtOH to achieve similar signal strength. Prior to each ionization mode a corona event of the opposite charge with a high voltage was necessary to get a stable and strong signal. Negative ionization consistently had roughly one order of magnitude less signal and was more susceptible to disturbances. Measurement of both modes in one run was only possible if the negative ionization preceded the positive ionization. Residual proteins, lipids and other analytes started to build-up on the needle (see Supplementary Figure 1) after 0.5 to 1 min and signal intensity started to notably decrease after 2 min. Signal clarity during the search for optimal voltage and needle immersion depths profited from needle changes for each new setting and short measurements <2 min.

**Supplementary Figure 1:**
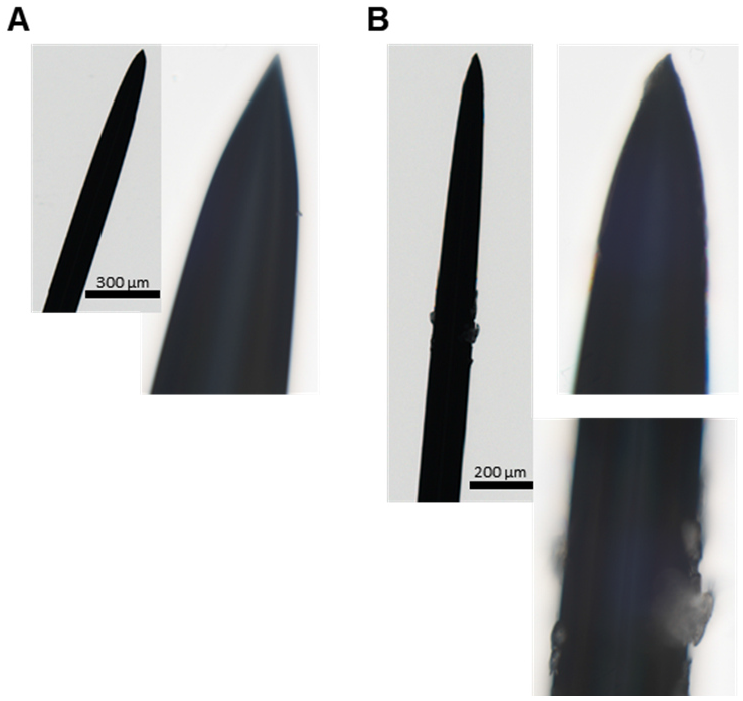
Microscopy images of (A) fresh needle and (B) a needle after 1 min measurement in negative ionization mode. Microscopy images were captured with 40x magnification in brightfield with a Vectra3 System from Akoya Biosciences (software was Vectra 3.0). Image sections were extracted and exported as TIFFs with InForm 2.4.

Finally, a 1:20 dilution with 10 mM NH_4_Ac 70% MeOH 5% DMSO was selected over 10 mM NH_4_Ac 50% EtOH 5% DMSO for two reasons: (1) MeOH had a more stable negative ion mode, (2) MeOH precipitates were removable with AcroPrep 0.2 µm PTFE vacuum filters (Pall Corporation, Ref 8047, AcroPrep Advance 96 Well 350µl 0.2µm PTFE Short Tip Natural PP), while EtOH clogged the filters. The compatibility to vacuum or positive pressure filtrations is crucial for later automatization, which itself is crucial for high-throughput in routine clinical laboratories or biobanks.

Extract supernatants become unstable within the first half hour at room temperature and after 2 h in a water ice bath (simulating a 4°C cooled auto-sampler). Therefore, either measurements should be done directly after preparation or extracts should be stored at -80°C until measurement, as done here. During the measurement of the 100 samples from the individuals, 13.6% of the measurements had invalid TIC patterns in either one or both ionization modes (see Supplementary Figure 3, 1. step). Likely, there is further potential for optimization of the established sample preparation method or instrument settings or set-up (e.g. needle material, auto-sampler).

## Supplementary Figures

**Supplementary Figure 2:**
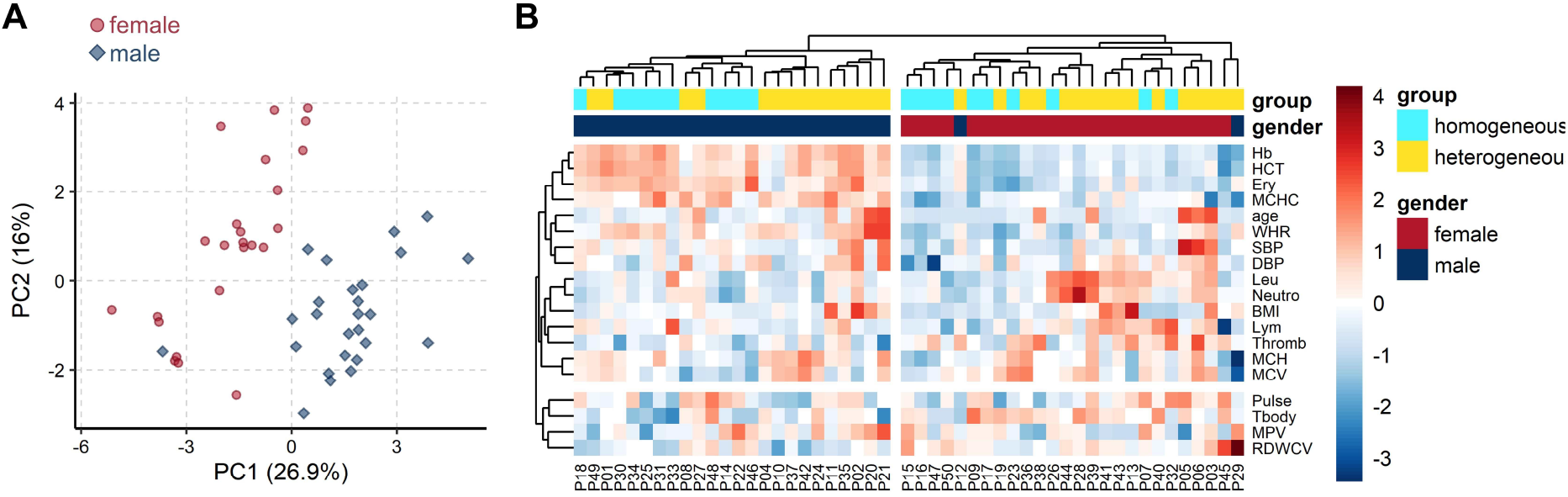
Routine hematological and clinical parameters were normal and differed between females and males. **A:** In the unsupervised PCA scores plot no strong outliers were detected, verifying biological inclusion of all 50 volunteers. No separation according to the study subgroups (homo-/heterogeneous) were detected. **B:** Heatmaps with hierarchical clustering underline the clear difference between genders and the absence of a systematic difference according to study subgroup homo- or heterogeneous.

**Supplementary Figure 3:**
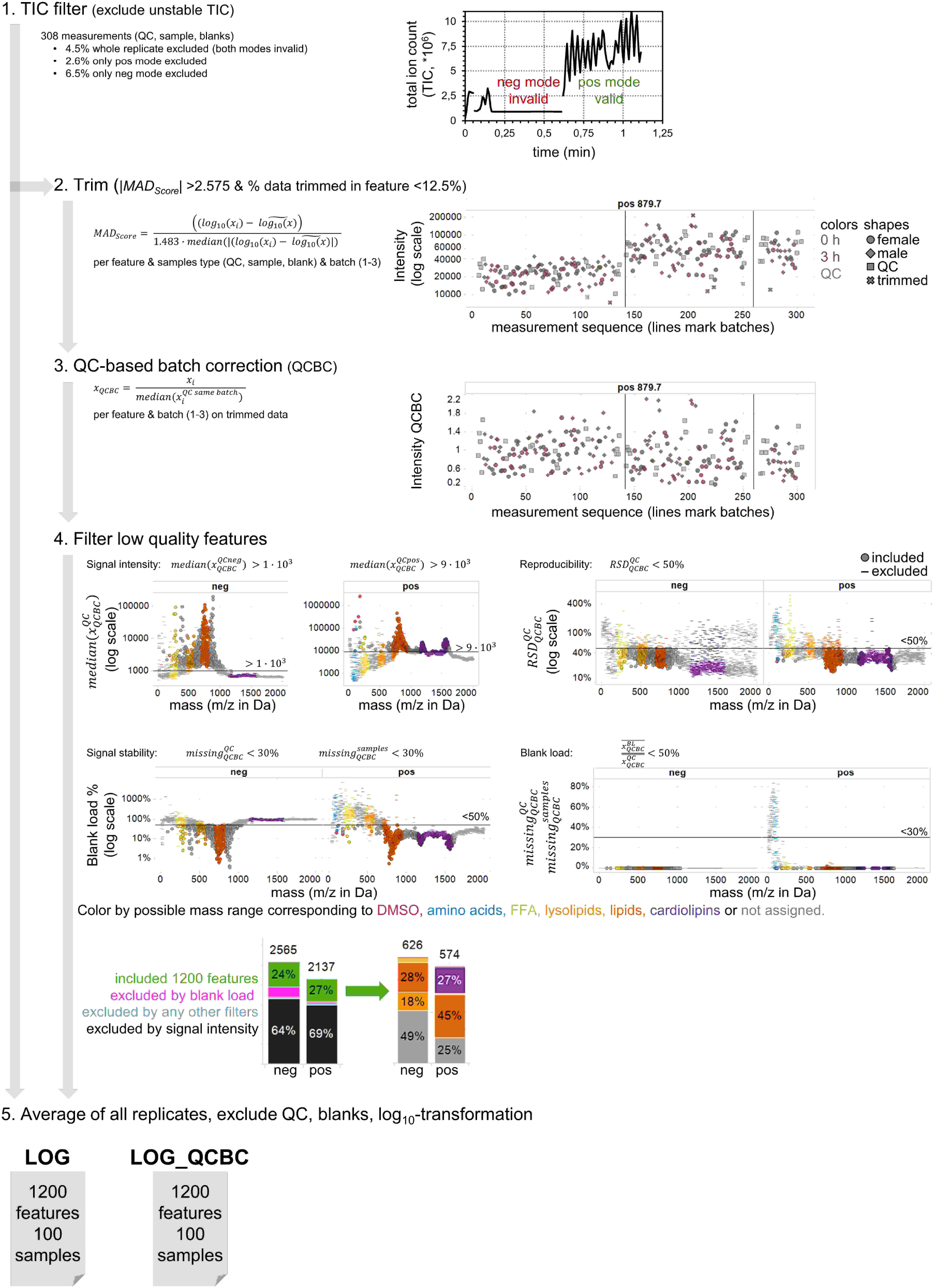
Overview of data extraction and pre-processing steps as described in Materials & Methods Table 4. Example plots are shown beside the steps. **1. Step** - TIC filter: Example of a single mode, invalid TIC pattern marked for exclusion (from Fig. 1). **2. Step** - Trim: Data trimming was based on MAD score, an outlier robust metric. Excluded single values are highlighted as crosses. The batch effect is well visible as intensity jumps between the 3 different measurement days. **3. Step** - QC-based batch correction: The robust and simple QC-based approach of normalizing all values in one batch to the median of the QCs in that batch was used and minimized batch effect sufficiently. **4. Step** - Filter low quality features: Four different markers of low analytical quality were applied. Most features were excluded due to very low signal intensity and stemmed mostly from baseline noise. Notably more features were excluded in neg mode due to high blank loads. Comparable feature numbers were found in both ionization modes before and after filtration. Possible metabolite classes based on mass ranges were added for a tentative orientation: amino acids 73–119 m/z FFA 198–331 m/z, lysolipids 469–594 m/z, lipids 594–975 m/z, cardiolipins 1150–1600 m/z. **5. Step** - Average of all replicates, exclude QC, blanks and apply log_10_-transformation to improve data distribution. Two data types were created, LOG and LOG_QCBC. For both the same filtered 1200 features were used for further analysis.

**Supplementary Figure 4:**
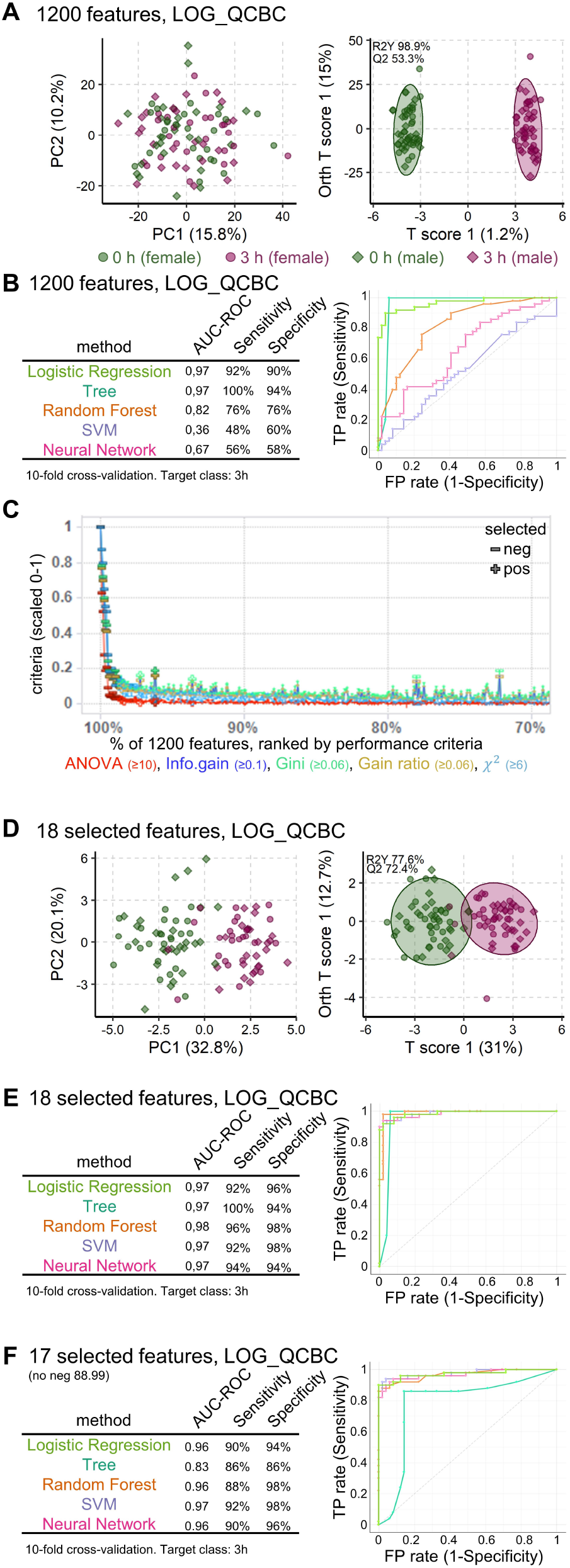
Plasma preparation delay is detectable from selected PESI features with high specificity in LOG_QCBC data. **A:** The unsupervised PCA (left) and supervised OPLS-DA (right) scores plot show no significant separation of the whole metabolome (1200 features, LOG_QCBC data from 50 volunteers) based on *time_delay* (0 h, 3 h). **B:** From five applied machine learning approaches only three were able to predict *time_delay* (AUC>0.8) and ROC curves differed notably between the different approaches. **C:** Feature importance in each of the five test criteria are shown ranked by highest importance first. The importance in each criterion was scaled 0-1 to allow direct comparison. For clarity only top 30% of data (360 features) are shown. The other features importance were below the inclusion threshold. **D:** Both PCA and OPLS-DA scores plot show a clear and highly significant difference for *time_delay* 0 h vs. 3 h, when based on the 18 most important features (LOG). **E:** All five applied machine learning algorithms delivered excellent predictions of *time_delay* (AUV>0.95) with no false negatives and very similar ROC curves.

**Supplementary Figure 5:**
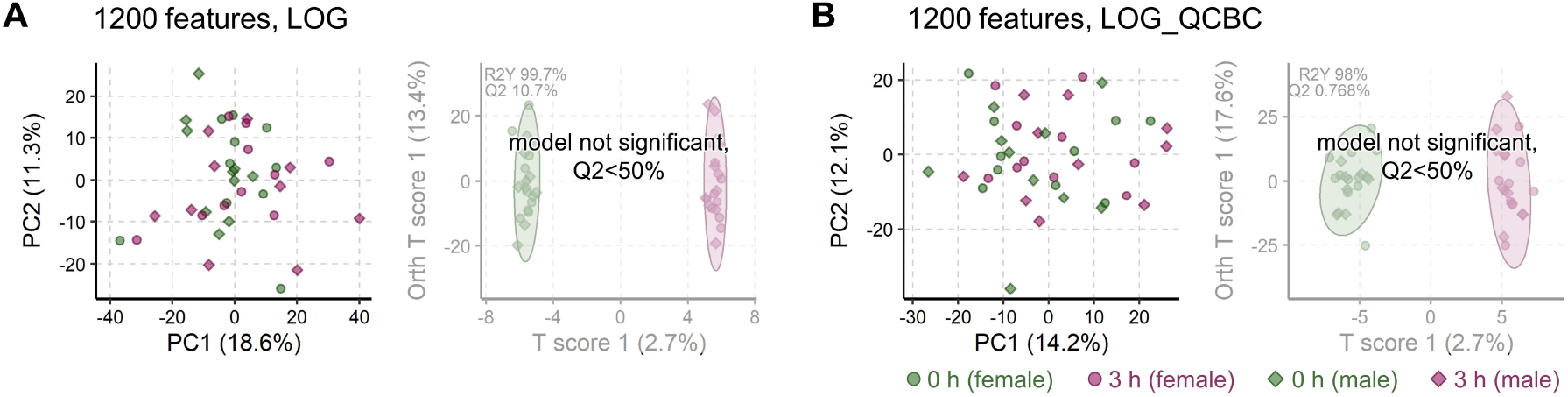
Concentration on homogeneous subgroup fails to improve time_delay separation in LOG or LOG_QCBC data with all 1200 features. **A/B:** The unsupervised PCA (left in panel) and supervised OPLS-DA (right in panel) scores plot show no significant separation of the whole metabolome in the homogeneous group (n=20) based on *time_delay* (0 h, 3 h) in both data types LOG (panel **A**) and LOG_QCBC (panel **B**).

**Supplementary Figure 6:**
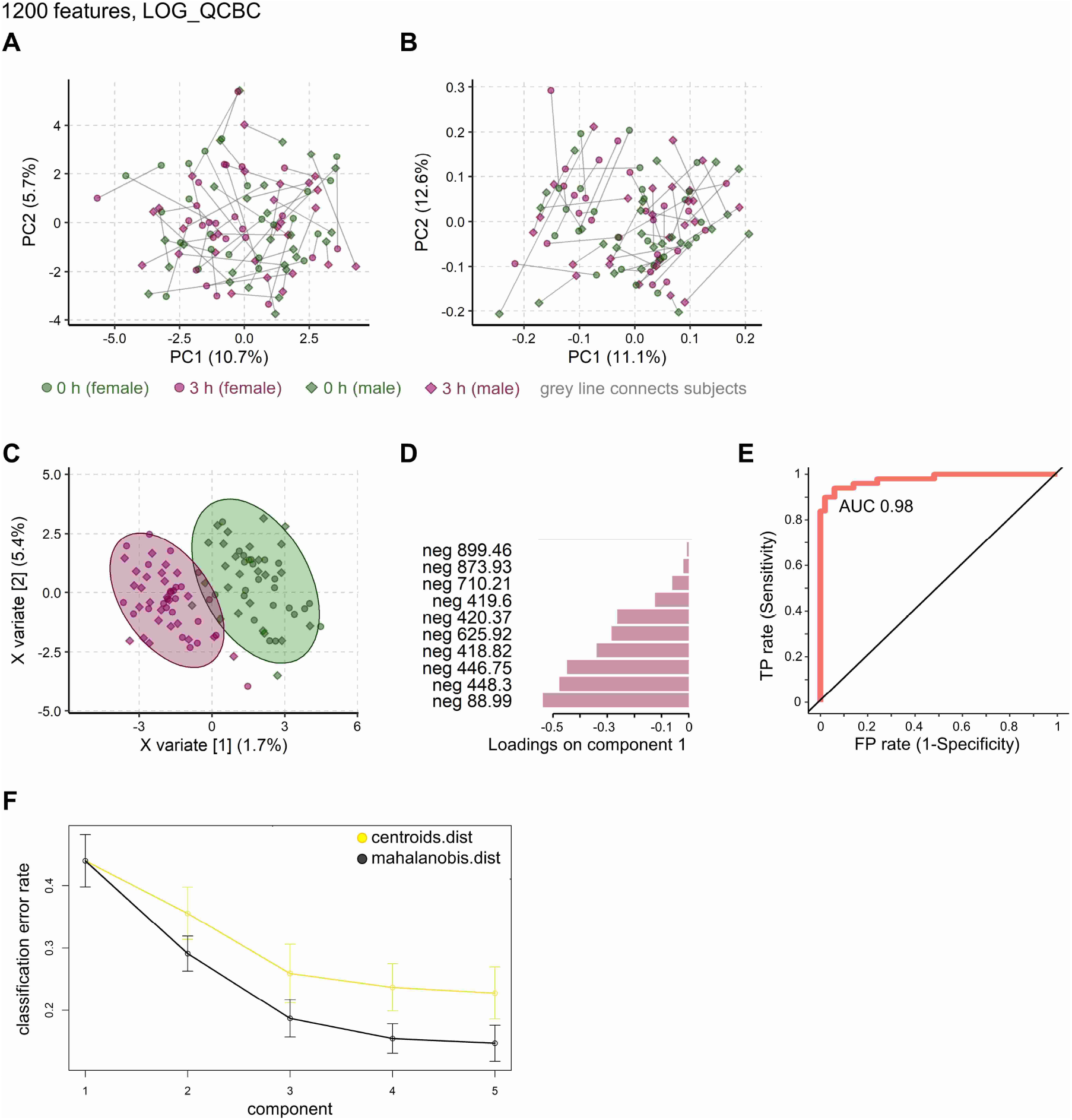
Sparse PCA and independent PCA fail to improve *time_delay* separation, while sparse PLS-DA finds significant differences. Analyses were based on LOG_QCBC, samples from all 50 volunteers (*time_delay* 0 h and 3 h) and all 1200 features. Sparse methods were set to keep at two components, 10 features in first component and 5 in second component. **A:** The scores plot of the sparse PCA shows no specific group separation according to *time_delay*. **B:** Analog, independent PCA shows no group separation by *time_delay*. **C:** Global sample plot of sparse PLS-DA with confidence ellipse shows a separation according to *time_delay*. **D:** The in C detected separation is driven by the 10 selected features. Features are colored by highest group mean (here all 3 h). **E:** ROC curve for 3 h vs. 0 h with first component. **F:** Performance per component (BER) in classification for two prediction distances (7-fold cross-validation, 50 repeats).

**Supplementary Figure 7:**
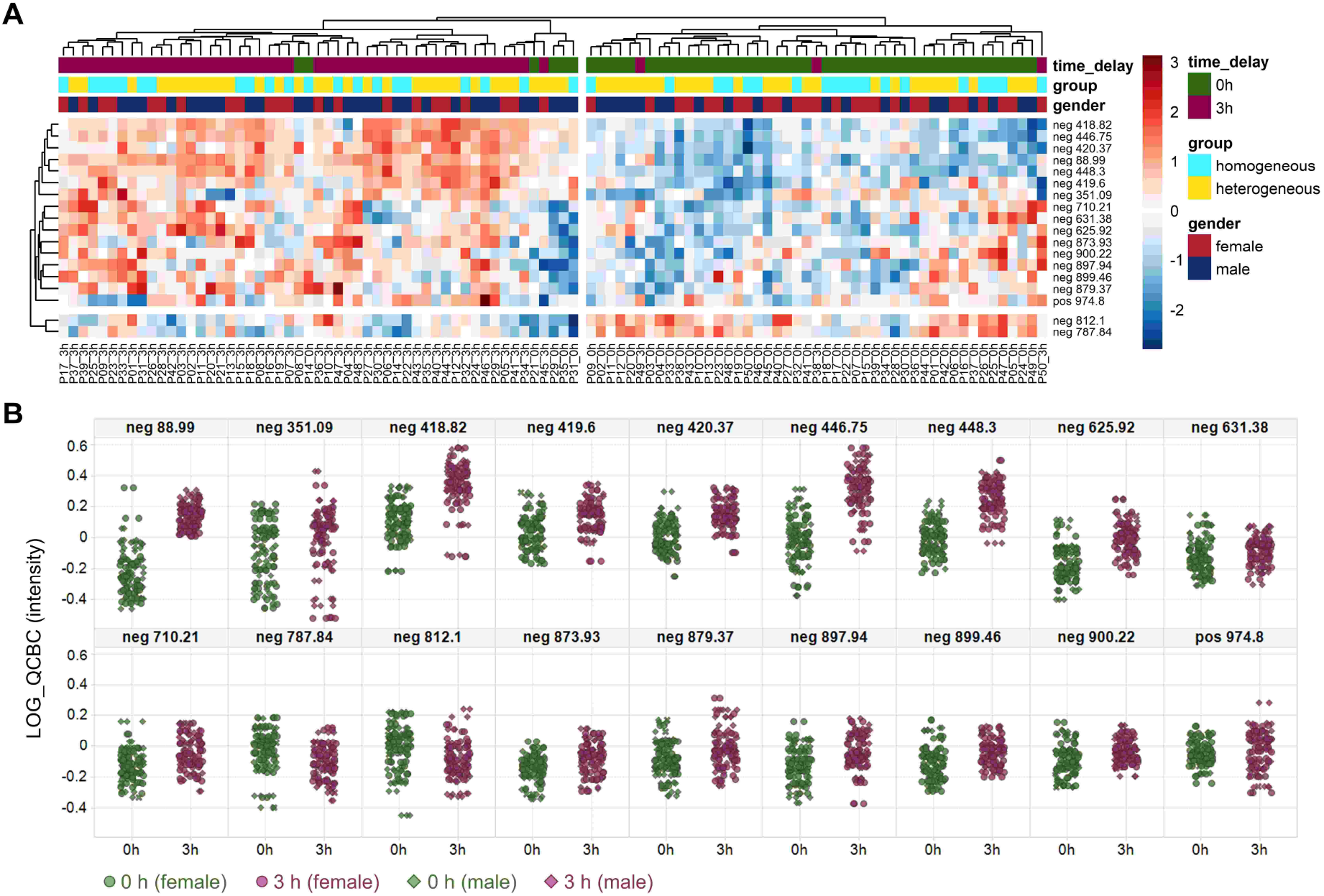
Heatmaps and plots of the 18 selected features in LOG_QCBC data. **A:** Heatmaps with hierarchical clustering underline the clear difference induced by *time_delay* in the 18 most important features. There are no systematic differences between study subgroups or genders. **B:** Scatter plots showing that from the 18 features, most (16) increased after the 3 h *time_delay* while only two feature levels decreased.

